# Ontogeny of plasma lipid metabolism in pregnancy and early childhood: a longitudinal population study

**DOI:** 10.1101/2021.09.15.21263636

**Authors:** Satvika Burugupalli, Adam Alexander T. Smith, Gavriel Olshansky, Kevin Huynh, Corey Giles, Sudip Paul, Anh Nguyen, Thy Duong, Natalie Mellett, Michelle Cinel, Sartaj Ahmad Mir, Li Chen, Markus R. Wenk, Neerja Karnani, Fiona Collier, Richard Saffery, Peter Vuillermin, Anne-Louise Ponsonby, David Burgner, Peter J. Meikle, Barwon Infant Study Investigator team

## Abstract

**Background:** There is mounting evidence that *in utero* and early life exposures may predispose an individual to metabolic disorders in later life; and dysregulation of lipid metabolism is critical in such outcomes. However, there is limited knowledge about lipid metabolism and factors causing lipid dysregulation in early life that could result in adverse health outcomes in later life. In this study, we aim to understand the lipid metabolism in pregnancy, and from birth to four years.

**Methods and findings:** We performed comprehensive lipid profiling of 1074 mother-child dyads in the Barwon Infant Study (BIS), a population based pre-birth cohort and measured 776 distinct lipid species across 42 lipid classes using ultra high-performance liquid chromatography (UHPLC). We measured lipids in 1032 maternal serum samples at 28 weeks’ gestation, 893 cord serum samples at birth, 793, 735, and 511 plasma samples at six, twelve months, and four years, respectively. The lipidome differed between mother and newborn and changed markedly with increasing postnatal age. Cord serum was enriched with long chain poly-unsaturated fatty acids (LC-PUFAs), and corresponding cholesteryl esters relative to the maternal serum. Alkenyl-phosphatidylethanolamine species containing LC-PUFAs increased with postnatal age, whereas the corresponding lysophospholipids and triglycerides decreased. We performed regression analyses to investigate the associations of cord serum lipid species with birth factors: gestational age, birth weight, mode of birth and duration of labor. Majority of the cord serum lipids were strongly associated with gestational age and birth weight, with most lipids showing opposing associations. Each mode of birth showed an independent association with cord serum lipids.

**Conclusions:** There were marked changes in the plasma lipidome over the first four years of life. This study sheds light on lipid metabolism in infancy and early childhood and provide a framework to define the relationship between lipid metabolism and health outcomes in early childhood.

**Funding Statement:** This work was supported by the A*STAR-NHMRC joint call funding (1711624031).

## Introduction

The developmental origins of health and disease paradigm suggests that prenatal, perinatal, and postnatal heritable and environmental influences result in long-term developmental, physiological, and metabolic changes in major tissues and organs [1, 2]. These metabolic perturbations can contribute to later life disease risk, including cardiovascular diseases and related cardiometabolic conditions [3, 4]. Alterations to lipid metabolism are a key driver of metabolic disorders [5, 6]. The lipid profile in blood (the lipidome) provides an integrative measure of genetic and environmental exposures on circulating lipid metabolism and quantification of the lipidome offers a rapid and effective approach to identify biomarkers potentially predictive of several diseases [7]. Large population-based studies from our group and others have established specific lipids to be associated with cardiometabolic disorders including diabetes and cardiovascular diseases [8-11]. In contrast to adult data, little is known about lipid metabolism in early life. Understanding the key determinants of early life lipid metabolism will inform the development of risk-stratification and early interventions.

The Barwon Infant Study (BIS) is a longitudinal Australian birth cohort with antenatal recruitment and repeated in-depth data and sample collection that is designed to facilitate a detailed mechanistic investigation of disease development within an epidemiological framework [12]. In this study, we applied a comprehensive lipidomic approach to understand the lipid metabolism during pregnancy and throughout the first four years of life. We utilized targeted UHPLC to measure 776 distinct individual lipid species across 42 distinct lipid classes. This study provides baseline characterisation of the natural biological variation of circulating lipid species of the mother and the children at birth, six, twelve months and four years highlighting important lipid species associated with known metabolic disease risk factors.

## Methods

### Research design and the cohort

The Barwon Infant Study is a pre-birth cohort (n = 1,074 mother-infant pairs, 72% Anglo-European) from the Barwon region of south-eastern Australia [12]. Women were recruited prior to 28 weeks of pregnancy. Infants born prior to 32 weeks, who developed a serious illness in the first week of life, or who had significant congenital or genetic abnormalities, were excluded. Participants were reviewed at birth and at 1, 6, 9, 12, 24 and 48 months. Data on maternal age, birth order, prenatal weight, and antenatal comorbidities were collected from questionnaires and hospital records, and by standardized clinical examination. Cord blood was collected at birth in a serum clotting tube, samples were centrifuged within 2 h of collection and the serum was separated and stored at –80 °C. Maternal pre-pregnancy BMI was calculated from self-reported pre-pregnancy weight and directly measured maternal height at the first study visit (28–32-week gestation). Birth anthropometric measures (birthweight, length, and head circumference) were collected within the first 2 days of life. Anthropometric measurements were also obtained at birth, 6 months, 12 months and 4 years. BMI at each time point was calculated by weight (kilograms) / height (meters)^2^. Ethics approval for this study was granted by the Barwon Health Human Research and Ethics Committee (HREC 10/24).

### Lipidomic profiling

*Lipid extraction:* Lipid extraction was performed as described previously in Alshehry *et al*. [13]. In brief, 10μL of plasma was mixed with 100μL of butanol: methanol (1:1) with 10mM ammonium formate containing a mixture of internal standards. Samples were vortexed, sonicated for an hour and then centrifuged (14,000xg, 10 min, and 20 °C) before transferring the supernatant into sample vials with glass inserts for analysis.

#### Liquid chromatography-tandem mass spectrometry

Lipidomics was performed as described previously in [9] with adaptations for a dual column set up. Analysis of plasma extracts was performed on an Agilent 6490 QQQ mass spectrometer with an Agilent 1290 series HPLC system and two ZORBAX eclipse plus C18 column (2.1×100mm 1.8mm, Agilent) with the thermostat set at 45°C. Mass spectrometry analysis was performed in both positive and negative ion mode with dynamic scheduled multiple reaction monitoring (MRM). A detailed list of MRMs and internal standards in provided in Supplementary Table 15.

The running solvent consisted of solvent A: 50% H_2_O / 30% acetonitrile / 20% isopropanol (v/v/v) containing 10mM ammonium formate and solvent B: 1% H_2_O / 9% acetonitrile / 90% isopropanol (v/v/v) containing 10mM ammonium formate. To avoid peak tailing for acidic phospholipids, we passivized the instrument prior to each batch by running 0.5% phosphoric acid in 90% acetonitrile for 2 hours and subsequently flushing the system with 85 % H_2_O / 15 % acetonitrile prior to sample run.

We utilized a stepped linear gradient with a 12.9 minutes cycle time per sample and a 1μL sample injection. To increase throughput, we used a dual column set up to equilibrate the second column while the first is running a sample. The sample analytical gradient was as follows: starting with a flow rate of 0.4 mL/ minute at 15% B and increasing to 50% B over 2.5 minutes, then to 57% over 0.1 minutes, to 70% over 6.4 minutes, to 93% over 0.1 minute, to 96% over 1.9 minutes and finally to 100% over 0.1 minute. The solvent was then held at 100% B for 0.9 minutes (total 12.0 minutes). Equilibration was started as follows: solvent was decreased from 100% B to 15% B over 0.2 minutes and held for an additional 0.8 minutes for a total run time of 12.9 minutes per sample. The next sample is injected, and the columns are switched.

The following mass spectrometer conditions were used: gas temperature, 150°C, gas flow rate 17 L/ min, nebulizer 20 psi, sheath gas temperature 200 °C, capillary voltage 3500 V and sheath gas flow 10 L/ min. Isolation widths for Q1 and Q3 were set to “unit” resolution (0.7 amu).

Plasma QC (PQC) samples consisting of a pooled set of plasma samples taken from 6 healthy individuals and extracted alongside the study samples were incorporated into the analysis at 1 PQC per 20 plasma samples. Technical QC samples (TQC) consisted of PQC extracts which were pooled, then split into individual vials to provide a measure of technical variation from the mass spectrometer only. These were included at a ratio of 1 TQC per 20 plasma samples. TQCs were monitored for changes in peak area, width, and retention time to determine the performance of the LC-MS/ MS analysis and were subsequently used to account for differential responses across the analytical batches. To align the results to any future datasets, the NIST 1950 SRM sample (Sigma) was included as a reference plasma sample, at a rate of 1 per 40 samples.

Relative quantification of lipid species was determined by comparison to the relevant internal standard. Lipid class total concentrations were calculated as the sum of individual lipid species concentrations, except in the case of triacylglycerides (TGs) and alkyl-diacylglycerides (TG-Os), where we measured both neutral loss [NL] and single ion monitoring [SIM] peaks, and subsequently used the abundant but less structurally resolved [SIM] species concentrations for summation purposes when examining lipid class totals.

#### Data processing

Lipidomic analysis was performed across 11 batches of ∼500 samples with quality control samples analysed every 20 samples. The chromatographic peaks were integrated using the Mass Hunter (B.07.00, Agilent Technologies) software and assigned to specific lipid species based on MRM (precursor/product) ion pairs and retention time. Upon completion of quantification, lipidome data for 777 lipid features in 4007 samples was available across 11 batches. Zero values (values beneath the mass spectrometer’s detection threshold) were replaced by 1/10^th^ the minimum value for the concerned lipids in the corresponding sample types. We then aligned the lipid concentrations across different batches by median centring the PQC quality control samples, and additionally scaled all batch-wise standard deviations for biological samples (on the log scale) to the mean standard deviation. We visually assessed lipids for large-scale technical variations (such as groups of outlier measurements, effects of drift or lipid hydrolysis over batch run duration), and re-quantified affected lipids where possible, and excluding them or selectively setting some values to missing where not. Missing values were then imputed using a k-nearest neighbour approach in the samples space. We used two different approaches for outlier sample detection. On one hand, lipid concentrations were log-transformed, and Z-scored, and absolute Z-scores were summed for each sample as a measure of their extremeness. Samples showing sums above the 95 percentile were flagged as potential outliers. On the other hand, the Z-scores were used in a Principal Components Analysis (PCA), and the 7 first principal components were retained as explaining the most variance. In the space defined by these components, each sample’s distance to the origin was calculated as a measure of the sample’s extremeness. Samples with a distance greater than the 95%-ile were flagged as potential outliers. Samples flagged by both approaches were declared outliers are removed from further analysis. Further inspection revealed that 16 samples were “missed injections” on the mass spectrometer, 27 cord serum samples were contaminated by maternal blood, these which were excluded, leaving 3964 samples in total. Ultimately, we retained 776 lipid measures across all batches. SIM-based lipid measures were not included in lipid species-level analyses, leaving 733 species.

### Statistical analysis

All statistical analyses were carried out in R (3.5.0 or 4.0.3). Plasma and cord serum lipid concentrations were log10 transformed prior to statistical analysis. PCA was performed using package FactoMineR using default settings.

Associations between participant characteristics (age, sex, BMI) and lipid species were determined using multiple linear regression, adjusting for appropriate covariates in each analysis [14]. All the analyses at birth using cord serum were adjusted for birth weight, gestational age, mode of birth, duration of labor, sex, maternal pre-pregnancy BMI, maternal gestational diabetes mellitus (GDM), maternal education, maternal age, birth order and lipidomics run batch.

Longitudinal analyses (associations of lipid species with sex, BMI and weight across infancy and early childhood) were performed using linear regression models of sex/ BMI/ weight against the log-transformed abundance of lipid species at each time point, adjusted for sex and age. We observed that gestational age influences lipids at 6 months and not at 12 months and hence adjusted for gestational age at the 6 month time point. Additionally, the 6 and 12 month analyses were adjusted for breastfeeding status, dichotomised at each time point as “currently/not currently breastfeeding” based on breastfeeding questionnaire data. The effective sample size varied slightly with each time point and between models due to missing samples or missing values for some covariates. Beta coefficients and 95% confidence intervals were then converted to percentage difference (percentage change = (10^β-coefficient – 1) x 100) to facilitate interpretation of results. All p-values were corrected for multiple comparisons, per model term, using the method of Benjamini and Hochberg [15]. Differences between the maternal antenatal plasma lipidome and the cord serum lipidome were tested using paired Wilcoxon Ranked Sum tests.

## Results

The baseline characteristics of the complete cohort are shown in Supplementary Table 1. In this study, we utilized serum samples of the mothers at 28 weeks’ gestation, cord serum samples at birth, and plasma samples at six, twelve months, and four years.

### Serum/Plasma lipidomics

We measured the plasma lipid species in a total of 3964 samples: 1032 in mothers, 893 at birth, 793 at six months, 735 at twelve months, 511 at four years (Fig 1a). Of the measured lipid species and classes, we retained 733 lipid species across 37 classes for further analyses. The median coefficient of variation (CV) for the lipid measures based on the plasma quality control (PQC) samples was 10.1%, with 92.6% of lipid measures showing a CV <20%. **The serum/plasma lipidome differs between mothers, newborns, and infants**. Overall lipid levels increased with age: maternal serum had higher total lipid levels than infant plasma, which in turn had higher levels than cord serum (Fig 1d). Principal Component Analysis (PCA) of the lipidomic data from all participants at all the time points revealed a clear separation of maternal, newborn, and infant samples across the first and second principal components (Fig 1b). A PCA of the infant samples showed further separation of the 6, 12- and 4-year time points (Fig 1c).

**Fig. 1.**
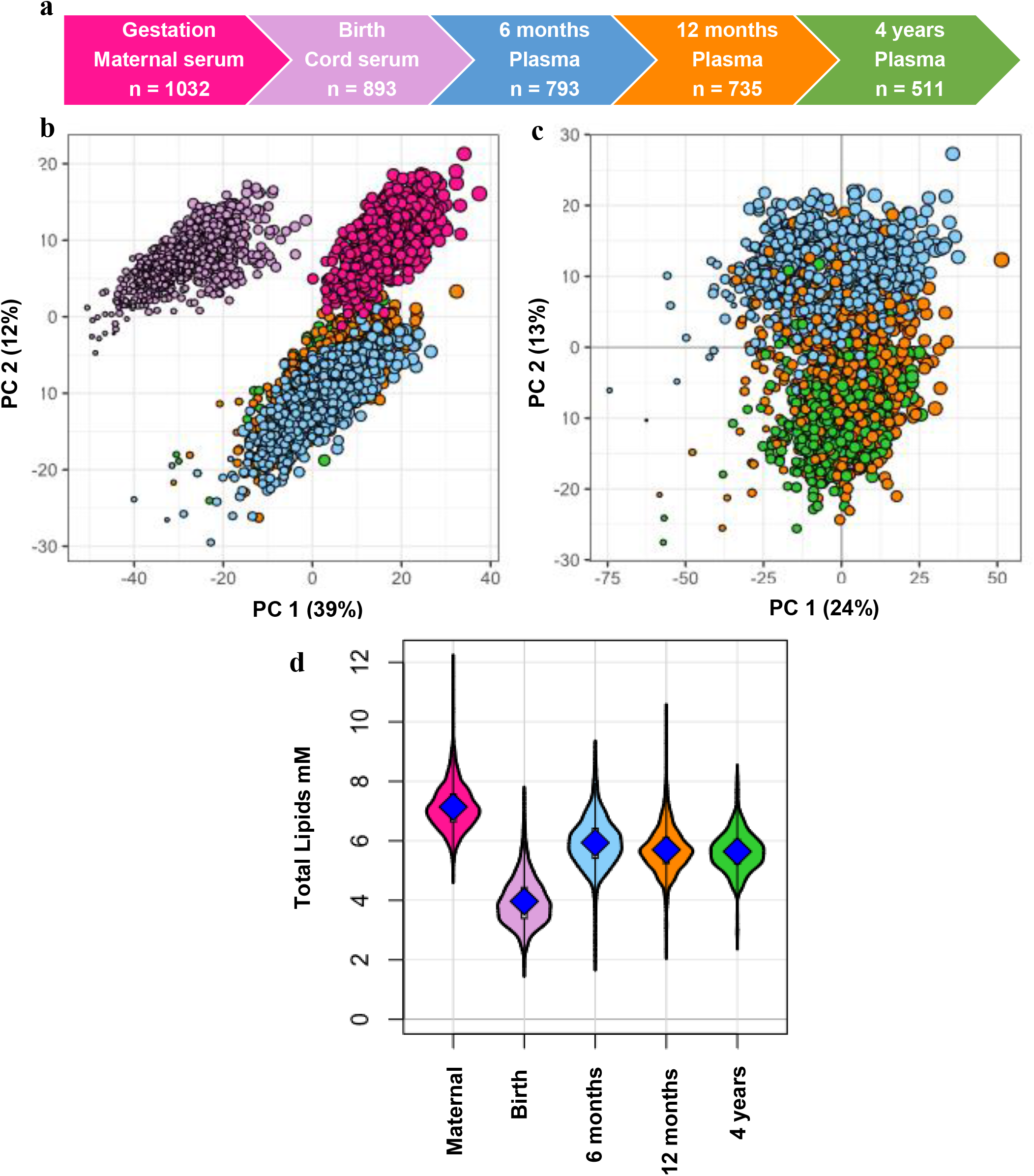
Snapshot of the Barwon Infant Study plasma lipidomics. **a,** The BIS timeline and final lipidomic sample numbers at each time point. **b**, PCA of the lipidomic measures at all the time points, marker size is proportional to the median log total lipid level (%) **c**, PCA of the lipidomic measures at 6 months, 12 months and 4 years, marker size as previously stated. **d**, Plasma total lipid concentration in maternal and infant sample groups. For all panels: Maternal serum, M28, pink colour; newborn cord serum, birth, purple colour; 6-month plasma, blue colour; infant 12-month plasma, orange colour; 4-year plasma, green colour.

### Gestational age and birthweight influence newborn cord serum lipids

461 (of 733, 63%) cord lipid species were associated with gestational age and 299 (of 733, 41%) lipid species were associated with birth weight. Gestational age was modestly correlated with birth weight (r = 0.45). However, the lipid profile associated with birth weight was discordant with the lipid profile associated with gestational age as indicated by the plot of the beta coefficients for gestational age against the beta coefficients for birth weight (y = -10.535x + 0.0101, r^2^ = 0.714, Supplementary Fig. 1). 591 (of 733, 80%) lipid species had an opposing association with gestational and birth weight. Species of di- and tri-acylglycerol, alkyl-diacylglycerol, acylcarnitine and free fatty acids (FFA) increased with gestational age but decreased with birth weight. Phospholipid species containing either an odd-numbered straight, or a methyl branched fatty acid (15-methylhexadecanoic acid, 15-MHDA) were positively associated with gestational age (Supplementary Fig. 2a) but negatively associated with birth weight (Supplementary Fig. 2b). While lysophospholipid species as a class were negatively associated with gestational age, we observed elevated levels of odd and branched chain fatty acid containing lysophosphatidylcholine species: LPC (19:0) [sn1] and [sn2], LPC (15-MHDA) [sn1] and [sn2] with gestational age that had an opposite association with birth weight. Cholesteryl esters (CE) also showed a similar trend as seen by elevated levels of CE (15:0) and CE (17:0). Alkyl and alkenyl phosphatidylethanolamine species increased with both gestational age and birth weight. Species of lysophospholipids, alkyl and alkenyl phosphatidylcholine, dihydroceramide, hexosylceramide, cholesteryl ester and dehydrocholesteryl ester decreased with gestational age and increased with birth weight.

### Mode of birth and duration of labor are associated with specific signatures in the cord serum lipidome

In this study, we compared four different birth modes: assisted vaginal birth (assisted VB) (including both forceps- and vacuum-assisted birth), scheduled cesarean (CS) and unscheduled cesarean (unscheduled CS), with unassisted vaginal birth (VB) as reference. 214 lipid species (of 733, 29%) showed evidence of association with at least one mode of birth (Fig. 3). Lipid species of acylcarnitine, di- and triacylglycerols were elevated in both assisted VB and unscheduled CS relative to VB, while lipid species of free fatty acids, lysophosphatidylcholine decreased in both CS and unscheduled CS birth modes compared to VB (Fig. 3a, b, c, and Supplementary table 4 and 5). Unscheduled CS birth had an intermediate profile where 74% of the lipid species showed a similar association to that of the lipid species associated with assisted VB (r^2^ = 0.543, Supplementary Fig. 3a), while 57% of the lipid species showed the same trend as seen in lipid species associated with a CS birth mode.

**Fig 2.**
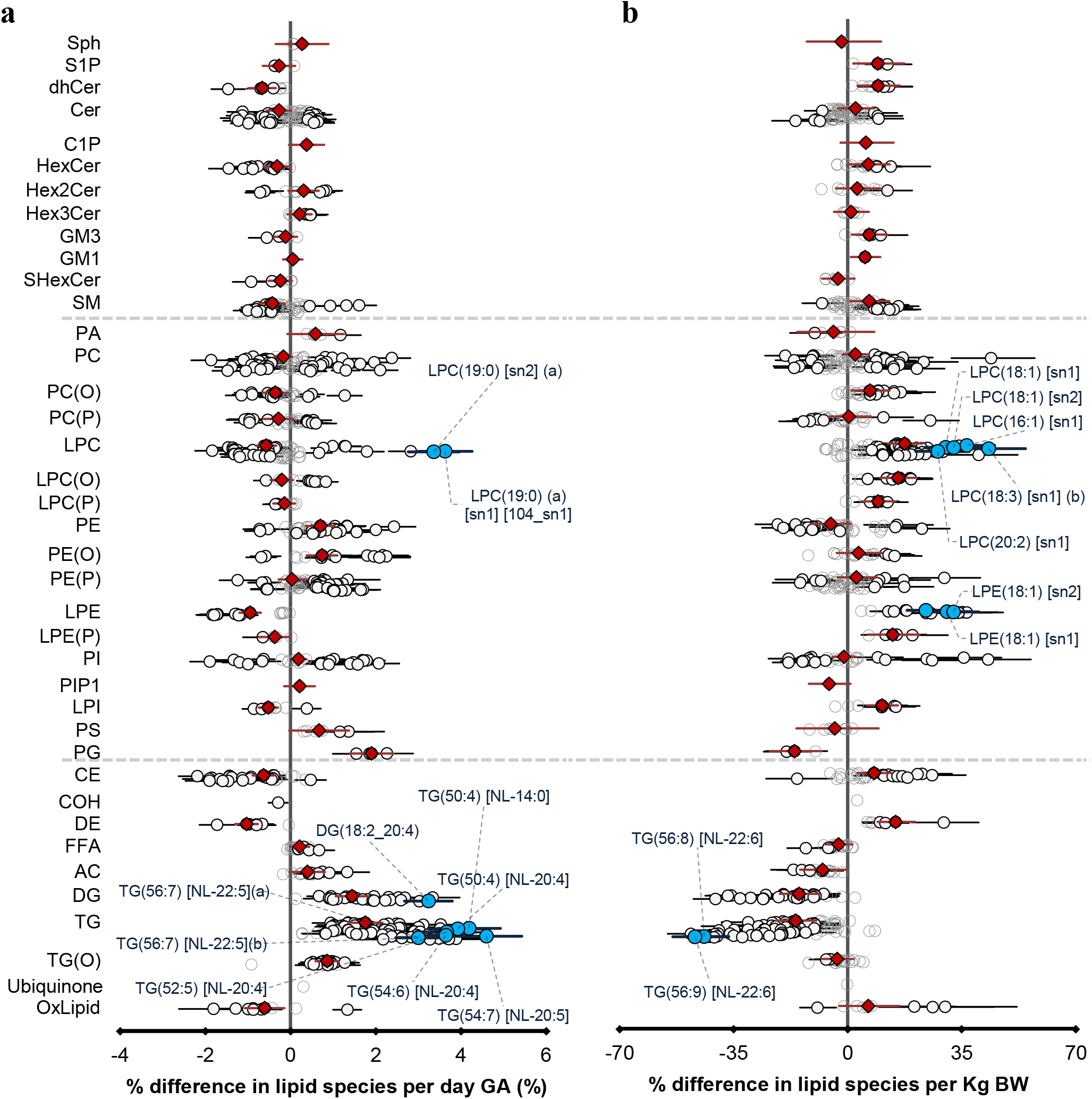
The newborn cord serum lipidome is influenced by gestational age (GA) and birth weight (BW). **a,** Estimated percentage difference of the lipid species per day increase in gestational age, determined using linear regression, adjusted for weight, sex, mode of birth, duration of labor, maternal pre-pregnancy BMI, GDM, maternal education, birth order and lipidomics run batch. **b**, Estimated percentage difference of the individual lipid species per kilogram increase in birthweight, determined using the same model as **a**. Grey open circles show non-significant species, white closed circles show significant lipid species (p < 0.05), the top 10 lipid species are shown in blue, red diamonds show lipid classes. Corrected p-value < 2.88E^-24^ and 1.81E^-15^, respectively. All p-values were corrected for multiple comparisons. Error bars represent 95% confidence intervals.

**Fig 3.**
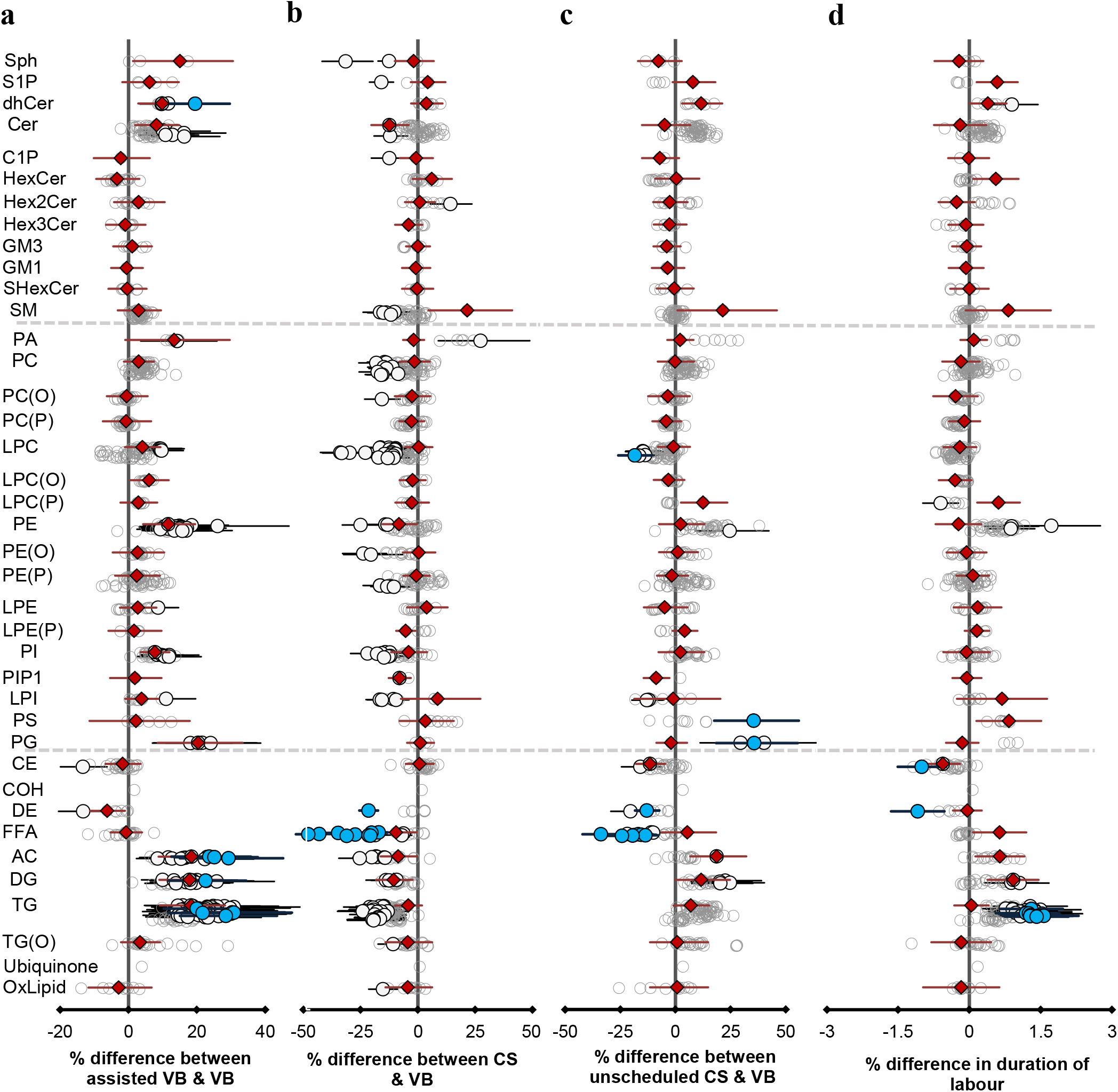
Mode of birth influences the newborn cord serum lipidome. Linear regression of the lipidome on delivery mode, each mode: assisted vaginal birth (assisted VB), scheduled cesarean (CS), unscheduled cesarean (Unscheduled CS) being compared to unassisted vaginal birth (VB), adjusting for birth weight, gestational age, sex, duration of labor, maternal pre-pregnancy BMI, GDM, maternal education, birth order and lipidomics run batch. Results are shown as % difference in lipids between each mode and VB. **a**, Lipid species associated with assisted vaginal delivery. **b**, Lipid species associated with scheduled cesarean delivery. **c**, Lipid species associated with unscheduled cesarean delivery. **d**, Lipid species associated with duration of labor, independent of the mode of delivery. Grey open circles show non-significant species, white closed circles show significant lipid species (p < 0.05), the top ten lipid species are shown in blue, red diamonds represent lipid classes. All p-values are corrected for multiple testing comparison. Error bars represent 95% confidence intervals. Corrected p value < 1.64E^-03^, 2.16E^-12^, 2.27E^-03^, 1.8E^-02^, respectively.

26 lipid species (of 733, 4%) showed evidence of association with duration of labor independent of the mode of birth. Similar to assisted VB and unscheduled CS, we observed elevated levels of acylcarnitine, di and triacylglycerols with longer duration of labor.

### Long chain poly unsaturated fatty acids (LC-PUFAs) are enriched in newborn cord serum

We performed paired Wilcoxon tests to investigate the relationship between maternal and newborn lipidome. While 80% of the lipid species were significantly higher in the maternal serum, the lipid species of sphingosine and sphingosine-1-phosphate, phosphatidylserine, acylcarnitine, dehydrocholesteryl ester, glycosphingolipids and FFAs were higher in the newborn lipidome (Fig 4a). Omega-3 and omega-6 LC-PUFAs: arachidonic acid (20:4, n-6), eicosapentanoic acid (20:5, n-3), adrenic acid (22:4, n-6), docosapentaenoic acid (DPA, 22:5, n-6), docosahexaenoic acid (DHA, 22:6, n-3) were enriched in the cord serum (Fig. 5b, 5c). While lipid classes such as cholesteryl ester, lysophosphatidylcholine and triglycerides were higher in the maternal plasma individual lipid species containing LC-PUFAs were higher in cord serum. (Fig 4b, 4c, Supplementary Tables 6 and 7).

**Fig. 4.**
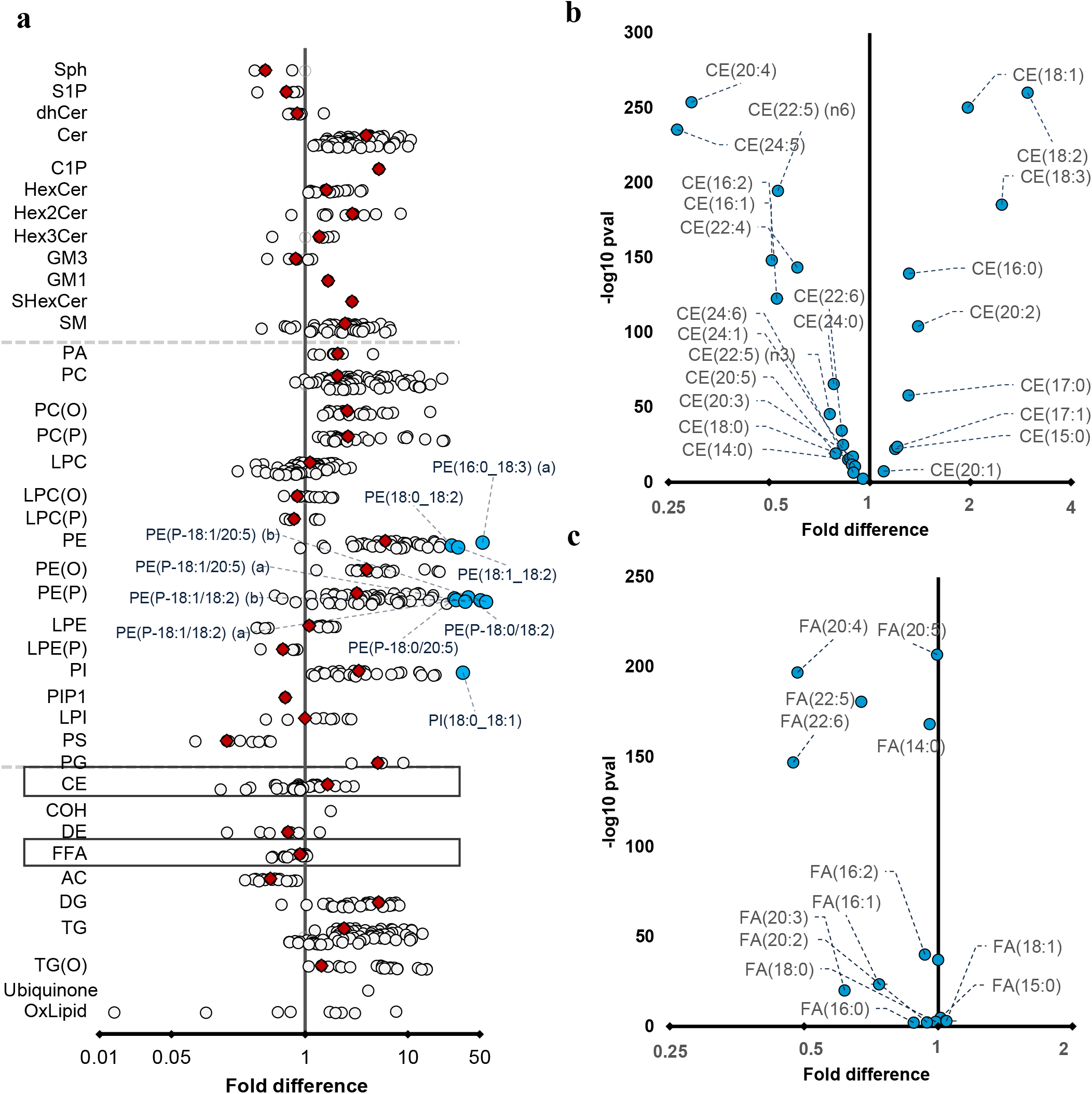
Differences in maternal and cord serum lipid species. Paired Wilcoxon tests were performed, and results are shown as fold differences of mothers relative to infants. **a**, Fold differences in lipid concentrations between maternal and cord serum. Cholesteryl esters and FFA classes are highlighted in boxes. **b**, Selective enrichment of cholesteryl esters containing long chain polyunsaturated fatty acids in cord serum. **c**, Selective enrichment of long chain polyunsaturated fatty acids in cord serum. Grey open circles show non-significant species, grey closed circles show significant lipid species (p < 0.05), the top 10 lipid species are shown in blue in panel a (corrected p value < 5.06E^-261^) and all significant lipid species are shown in blue in panel b and c, red diamonds show lipid classes. All the p-values are after multiple testing comparison.

**Fig. 5.**
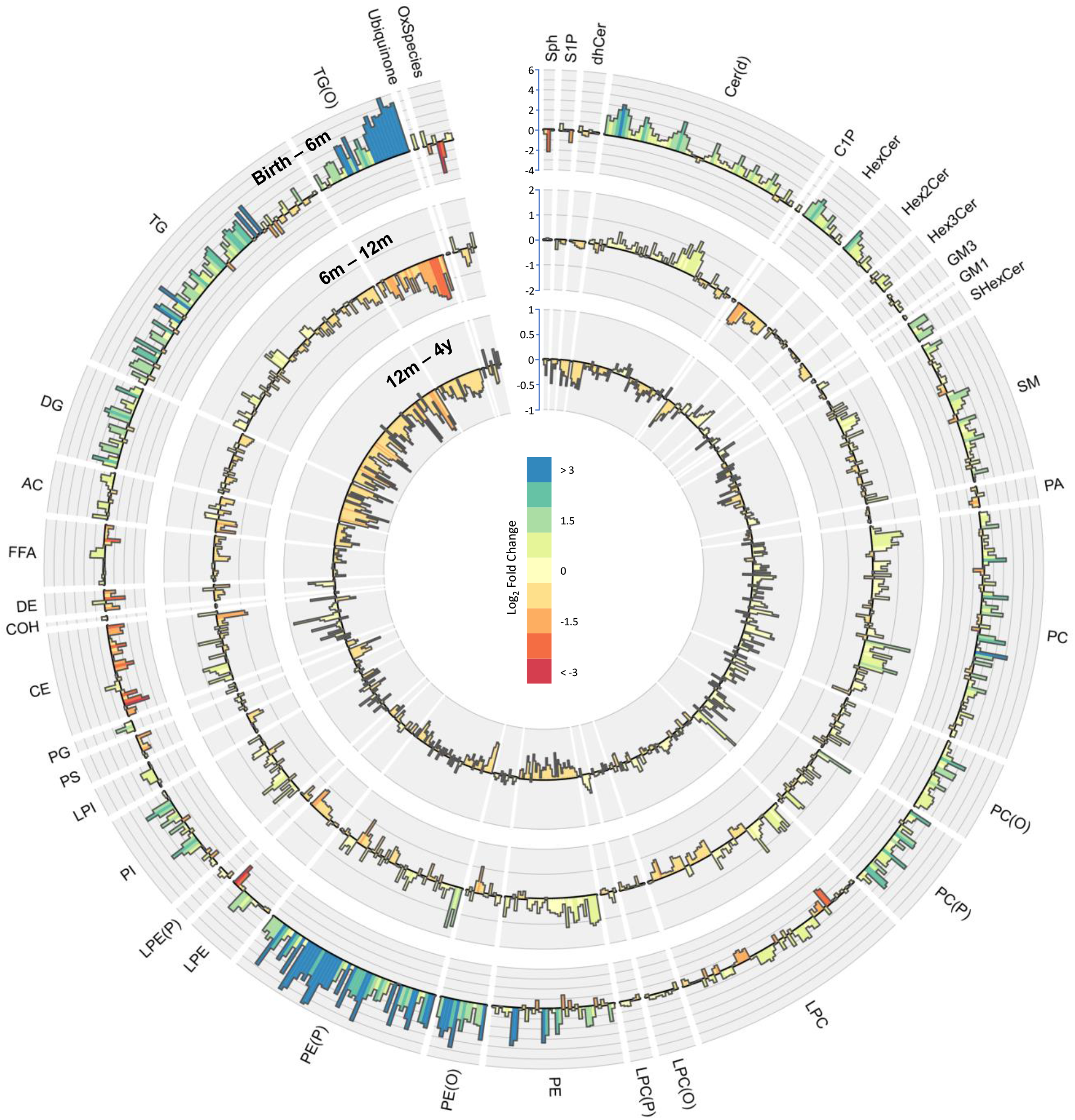
Lipidomic profile changes in the first four years. Paired t-tests were performed in infants with repeat measures (birth - 6 months n=646, 6 - 12 months n=628, 12 months – 4 years n=418), and results are shown as log2 fold change (FC). The outermost circle represents the changes in lipid concentration from birth to six months (axis range: -4 to 6), middle circle represents changes in lipid concentration six months to 12 months (axis range: -2 to 2) and the inner most circle represents the changes in lipid concentration from 12 months to four years (axis range: -1 to 1).

### The circulating lipidome changes over the first four years

We performed paired t-tests to study the changes in lipid concentration at different time points (Fig 5). 75% 549 (of 733, 75%) of the lipid species were higher at four years than at birth, with LC-PUFA containing alkenyl phosphatidylethanolamine species having the greatest increase. 123 lipid species (of 733, 17 %) consistently increased at all the time points from birth to four years, whereas 68 lipid species decreased across all the time points. There was a consistent decrease in LC-PUFA containing lysophospholipids and an increase in odd and branched chain fatty acids, and phospholipids containing these fatty acids, across all the time points.

#### Birth to six months

Majority of lipid species (558 of 733, 76%) were higher at six months compared to at birth. We observed the greatest increase in in species of alkenylphosphatidylethanolamine and alkyldiacylglycerol and the largest decrease in 20:4 and 20:6 containing lysophosphatidylcholine, lysophosphatidylethanolamine and cholesteryl esters (Supplementary Tables 8 and 9).

#### Six to twelve months

363 (of 733, 50 %) lipid species increased in concentration from six to twelve months. Odd and branched chain phospholipids showed the greatest increase while species of alkyl diacylglycerol and cholesteryl esters showed the greatest decrease (Supplementary Tables 8 and 9).

272 (of 733, 37%) lipid species consistently increased from birth to 6 months and from 6 to 12 months, while 90 (of 733, 12%) lipid species showed a consistent decrease. 365 (of 733, 50%) lipid species had an opposing trend at these time points. Lipid species of dihydroceramide, phospholipids, glycophospholipids, lysophospholipids, FFA, alkyl and alkenyl phospholipids, di- and tri-acylglycerols, acylcarnitine, sphingomyelin, mono and tri-hexosyl ceramides, ceramide-1-phosphate increased from birth to six months but decreased from six to twelve months. Lipid species of sphingosine decreased from birth to six months but increased from six to twelve months.

#### Twelve months to four years

284 (of 733, 37%) of the lipid species were higher at four years than at 12 months. We saw the highest increase in cholesteryl ester CE 20:5 and largest decrease in 22:6 containing di and triacylglycerols (Supplementary Tables 8 and 9).

Some lipid species showed variable changes as the child aged. Lipid species of dehydrocholesterol, alkenyl-lysophosphatidylcholine that decreased from birth to 6 months, and from 6 to 12 months, increased at four years. Lipid species of ceramide, phosphatidylethanolamine and phosphatidylglycerol that increased from birth to 6 months and 6 to 12 months, decreased at four years. Lipid species of sphingosine decreased from birth to 6 months, increased at 12 months, and decreased again at four years (Supplementary Fig 4).

Overall omega-3 and omega-6 LC-PUFAs: 20:3, 20:4, 22:4, 22:5 and 22:6 that were higher at birth, decreased by the time the infant reached six months, and continued decreasing further until four years. Most of the omega-3 PUFA cholesteryl esters: CE (22:5) (n-3) and 22:6 decreased in the first year but increased at 4 years except for CE (20:5) that decreased at 6 months but increased at 12 months and 4 years. Omega-6 PUFA cholesteryl esters: CE (20:4), CE (22:4) and CE (22:5) (n-6) decreased at 6 months but increased at 12 months and 4 years (Supplementary Table 10).

### Sex-specific changes in the circulating lipidome over the first four years

We investigated the influence of sex on lipid metabolism in early childhood. We found 218 lipid species (of 733, 29%) to be significantly associated at birth, 214 (of 733, 29%) at 6 months, 62 (of 733, 8%) at 12 months and 82 (of 733, 11%) at 4 years. We found significant differences in lipid species of ceramide, sphingomyelin, acylcarnitine, lysophosphatidylcholine, lysophosphatidylethanolamine, alkyl-lysophosphatidylcholine and triglycerides between boys and girls (Supplementary Tables 11, 12).

### The four-year lipid profile associated with BMI resembles an adult profile

Having shown that cord serum lipids are associated with birthweight, we then investigated the associations of circulating lipids with BMI and weight in infancy and at 4 years. At six months, 16 (of 733, 2%) lipid species were associated with weight and, 28 (of 733, 4%) with BMI; at 12 months 85 (of 733, 12%) lipid species were associated with weight and 13 (of 733, 2%) with BMI, and at four years 35 (of 733, 5%) lipid species were associated with weight and 1 with BMI (Supplementary Fig. 5, Supplementary Tables 13, 14, 15 and 16). At four years, the lipid profile associated with BMI more resembled that of an adult lipid profile associated with BMI. Lipid species of di- and tri-acylglycerols, sphingosine and sphingosine phosphate, glycosphingolipids, hexosylceramides, dihydroceramides, lysophospholipids, and the corresponding alkyl and alkenyl species were elevated with BMI similar to that seen in an adult profile. A scatter plot of the beta coefficients of lipid species associated with BMI at four years and pre-pregnancy BMI in mothers revealed the similarity between these two profiles as indicated by a positive slope (Supplementary Fig. 6a, 6b). Sphingomyelin species: SM (d18:0/14:0) and SM (d18:2/14:0) were significantly associated with weight across all time points and with BMI at birth, 6 months, and 12 months.

## Discussion

This is the first and largest study of the lipidome from birth and early childhood in a population-derived cohort. The lipidome was different between mother and child and changed markedly with postnatal age. We identified that gestational age and birth weight were associated with cord serum lipids. Most of the lipid associations with gestational age were in the opposing direction to the associations with birth weight. Mode of birth was also associated with the cord serum lipidome, and the profile differed from that observed for gestational age and birthweight. There were marked changes in the ontogeny of the plasma lipidome from birth to four years of age: LC-PUFA, and cholesteryl esters containing LC-PUFAs were enriched in cord serum relative to the maternal serum. LC-PUFA containing lipid species altered between birth and four years with an enrichment in alkenyl-phosphatidylethanolamine species and a reduction in lysophospholipids and triglycerides. Concentrations of free LC-PUFAs decreased at four years, but the corresponding cholesteryl esters increased.

In this study, we examined the association of gestational age, birthweight, and mode of birth with cord serum lipid concentrations. Di- and tri-acylglycerols were positively associated with gestational age, which may reflect an evolutionary adaptation in preparation for the metabolic stress of labor, a period of high energy expenditure [16] [17]. A concomitant increase in branched chain fatty acids and corresponding phospholipids, a downstream product of the mitochondrial catabolism of branched chain amino acids and elevated levels of acylcarnitines, both suggest increased mitochondrial activity in babies with greater gestational age [18]. Gestational age and birth weight showed overall inverse lipidomic profiles that were largely independent of each other. In babies with higher birth weight, the observed lower level of triglyceride levels could reflect better packaging within the adipose tissue vs free availability in the circulation for energy mobilization. These results have also been observed in an ethnically diverse cohort, Growing Up in Singapore Towards healthy Outcomes (GUSTO), revealing that these lipidome changes associated with birthweight are independent of ethnicity [19].

The lipid profile associated with gestational age resembles the lipid profile associated with BMI in adults. For example, in adults, acylcarnitines, di and tri-acylglycerols are positively associated with BMI and body weight [20] like that seen in babies with higher gestational age. One possible explanation is that these lipid classes are involved in energy mobilisation and hence more abundant in circulation thereby acting as a source of free fatty acids which are required for various biological pathways [21] [22]. Increasing epidemiological evidence suggests infants born late pre-term (34 – 36 weeks) and early term (37 – 38 weeks) are at higher risk of a host of adverse health outcomes including respiratory disorders, increased and prolonged hospitalizations and developmental delays [23-25]. Understanding the changes in lipid metabolism during the final stages of gestation may provide mechanistic insights into the increasingly recognised long-term health sequelae of preterm and late preterm births [26].

Numerous studies have found associations between mode of birth and child health outcomes, including risk of future overweight and obesity [27, 28]. Relative to assisted VB, VB is the most stressful for the foetus followed by unscheduled CS birth, while CS is the least stressful for the baby. Elevated levels of di- and tri-acylglycerols and acylcarnitine in assisted VB, unscheduled CS and prolonged labor indicate that stress *in utero* and during birth increases energy mobilization for high energy expenditure in these modes of birth [29-31]. Additionally, mode of birth also effects the neonate’s gut microbiota [32]. Newborns delivered vaginally show a higher diversity of bacteria than those born by CS [32, 33]. The specific lipidome signature associated with each mode of birth may reflect the differences in the gut microbiome, although whether this could manifest at the time of delivery is uncertain. Considering data suggesting an increased risk of metabolic disorders later in life associated with mode of birth [28, 34], it is important to understand differences in lipid metabolism by birth mode.

### Lipid differences in mother and newborn cord serum

Compared to the mother, we observed elevated levels of LC-PUFAs in cord serum which are critical for *in utero* brain development as well as the rapid brain development in the first year of life [35, 36]. The elevated levels of di- and tri-acylglycerols, and phospholipids observed in the maternal circulation most likely act a source of fatty acids for the foetus. Lipoprotein lipase and endothelial lipase in the syncytiotrophoblast hydrolyse circulating maternal triglycerides and phospholipids respectively to provide the pool of fatty acids for the fetus [37]. Free fatty acids in the syncytiotrophoblast basal membrane are transferred to foetal circulation directly through facilitated diffusion or using fatty acid carriers (FAT/CD36) and fatty acid binding proteins (FABPpm, FATP4). In the fetal circulation, these fatty acids then bind to a fetoprotein and are transported to the liver, where they are re-esterified into complex lipids and transported into circulation [38, 39].

LC-PUFA containing phosphatidylcholine species were also elevated in maternal serum. These phospholipids undergo a *sn*-1 cleavage by endothelial lipases in the syncytiotrophoblast producing LC-PUFA lysophosphatidylcholine species which are then transported across the placenta by the major facilitator superfamily domain containing 2A (MFSD2a) transporter proteins explaining the elevated levels of LC-PUFA containing LPCs in cord serum [40]. Elevated levels of LC-PUFA containing cholesteryl esters in the cord serum, might also be acting as an additional source of LC-PUFAs for the foetus.

Recent studies have shown altered placental function and thereby altered lipid transfer in mothers with gestational diabetes and higher pre-pregnancy BMI [41, 42]. However, long term ramifications of maternal metabolic conditions on offspring health are currently unknown. Our study contributes to the understanding of trans-placental transfer of specific lipid species. Further studies on how maternal metabolic conditions may impact this transfer may provide insight into perturbations in lipidome *in utero* and its long-term health consequences.

### Changes in lipid metabolism in the first four years of life

We observed pronounced changes in the lipid profiles in infancy and early childhood. Elevated levels of LC-PUFAs, cholesteryl esters and lysophospholipids seen in the cord serum started to diminish as the child reached six months of age. However, at 12 months and four years of age, we observed an increase in odd chain, essential fatty acids and LC-PUFA containing phospholipids, which could be acting as a source of PUFA. Recent studies have shown LC-PUFA deficiency in several childhood disorders such as asthma, cystic fibrosis, attention-deficit/hyperactivity disorders (ADHD), obesity and diabetes [35, 36]. A balanced intake of omega–6 and omega–3 fatty acids is essential for homeostasis and normal development throughout the life cycle. A diet rich in omega-6 LC-PUFAs can shift the metabolism to an atherogenic/ diabetic state [43]. On the other hand, the beneficial effects of omega-3 LC-PUFAs also depend on complex interaction between different nutrients, and polymorphism in genes involved in omega-3 fatty acid metabolism. Understanding the changes in lipidome early on in life provides a window of opportunity for early intervention and decrease the risk of future metabolic disorders.

### Effect of sex on lipids

There is evidence of sex-differences in lipidome from late gestation onwards [44]. For example, cord plasma concentrations of total cholesterol, HDL and LDL are higher in girls than in boys [45], in keeping with our findings. We have previously identified sphingomyelin SM (d18:2/14:0) as the key lipid differentiating sex in adult cohorts [8, 20]. At birth, we did not find SM (d18:2/14:0) to be different between male and female babies. However, at age 4 years, SM (d18:2/14:0) was significantly associated with sex and was lower in boys relative to girls (10.42% lower in boys, p = 5.53 E^-04^). In adults, the differential activity of the fatty acid desaturase (FADS3) is responsible for the elevated levels of SM (d18:2/14:0) in women [20, 46]. Our results suggest that FADS3 starts exhibiting this differential activity at four years. Additionally, sex differences in circulating gonadotropin levels during the first few months of life is well documented [47-49]. More recently, a differential hypothalamic-pituitary-adrenal (HPA) axis activity in response to fetal glucocorticoid exposure has been recorded between sexes at birth [50]. Understanding the sex differences in the lipidome at birth and early life could provide mechanistic insights into sex-differences in cardiometabolic diseases.

### Effect of BMI on infant lipid metabolism

We have previously reported the lipid profile pattern associated with BMI in healthy adults [8]. In the current study, at 4 years of age, the lipid profile associated with BMI closely resembled that of an adult, although the associations were less evident, suggesting that the ontogenesis of the lipidome is not yet complete at 4 years of age. Similar results have been reported in GUSTO, where they observed a 92% overlap of lipids associated with 6 years old BMI and lipids associated with postnatal maternal BMI [19].

### Strengths and Limitations

Major strengths of this study include repeated and extensive sampling in a large population-derived cohort at in pregnancy and early life with high quality, standardised meta-data.

The BIS cohort provides an opportunity to study the effects of environmental factors during early infancy. However, the lack of generalisability (less ethnically diverse, more affluent than the general population) may limit the understanding of outcomes in different ethnicities and socio-economic strata. It would be valuable to follow the children through adolescence to understand the long-term modulation of lipid metabolism and early markers of future health or disease outcomes. We are currently investigating these results in another large birth cohort, based in Singapore – the Growing Up in Singapore Towards a healthy Outcome (GUSTO) cohort to investigate whether similar changes are evident in a population of Asian ethnicity. Any lipid differences based on the matrix must be kept in mind as serum samples were used during gestation, and at birth vs plasma samples at other time points.

## Conclusion

We report in-depth lipidomic profiling in both gestation and the earliest stages of life, thereby providing insight into the ontogeny of lipid metabolism through infancy and early childhood. Despite the limitations, the extensive lipidomic profiling gives us insight into perinatal and postnatal factors associated with lipid metabolism during the first four years of life. There is now growing evidence of fetal programming emphasising the profound and sustained impact of intrauterine and early life factors to foetal health and development of metabolic diseases in later life. Understanding the baseline characteristics of lipid metabolism at birth and throughout early years provide a resource for further studies to elucidate the clinical implications. The datasets resulting from this study now provide a rich resource to further investigate the relationship between lipid metabolism and health outcomes during early life. Children within this study are continuing to be followed and future outcome data will build on these initial early life findings.

## Data Availability

Access to BIS data including all data used in this paper can be requested through the BIS Steering Committee by contacting the corresponding author. Requests to access cohort data are considered on scientific and ethical grounds and, if approved, provided under collaborative research agreements. Deidentified cohort data can be provided in Stata or CSV format. Additional project information, including cohort data description and access procedure, is available at the project's website https://www.barwoninfantstudy.org.au.

## Acknowledgements

The establishment work and infrastructure for the BIS was provided by the Murdoch Children’s Research Institute, Deakin University and Barwon Health. Funding has been provided by the National Health and Medical Research Council of Australia (607370, 1009044, 102997, 1082037, 1076667, and 1084017), the Jack Brockhoff Foundation, the Scobie Trust, the Shane O’Brien Memorial Asthma Foundation, the Our Women’s Our Children’s Fundraising Committee Barwon Health, the Rotary Club of Geelong, the Shepherd Foundation, the Ilhan Foundation, and the Operational Infrastructure Support Program of the Victorian Government. We acknowledge the participation and commitment of all the families in the BIS.

## Author Contribution

PJM conceived and supervised the study. DB, ALP, RS, and PV contributed to data and sample collection in BIS cohort. SB, AN, TD, NM, MC performed the sample sorting and lipidomics. SB, AAT performed the data analyses. GO, KH, CG contributed to data analysis and interpretation. SB, AAT, PJM interpreted the data and wrote the manuscript. All authors critically read and approved the manuscript content.

## Supplementary Figures

**Supplementary Fig 1.**
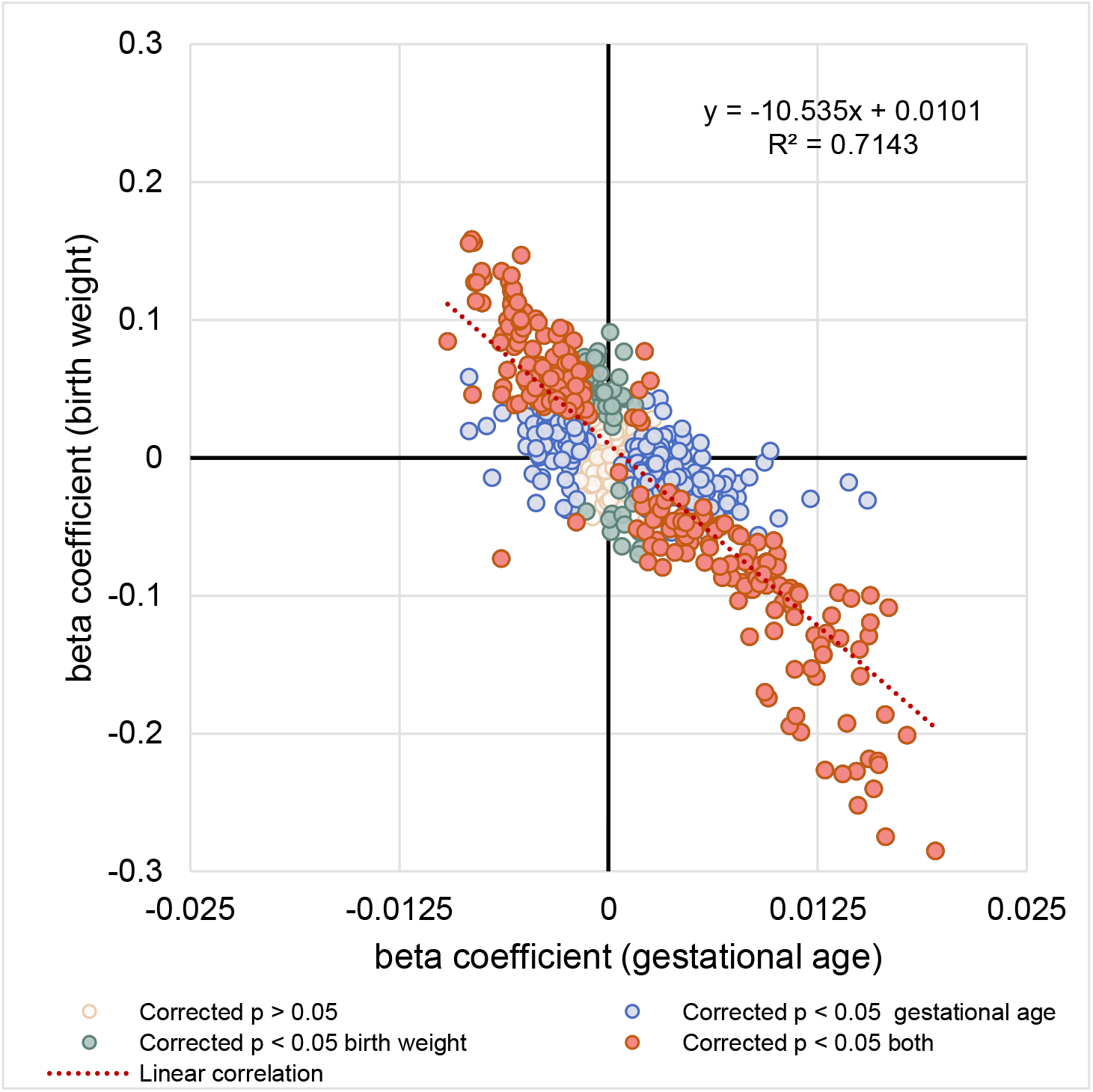
The correlation between regression coefficients of each lipid species associated with gestational age (x-axis) and lipid species associated with birth weight (y-axis) was examined.

**Supplementary Fig 2.**
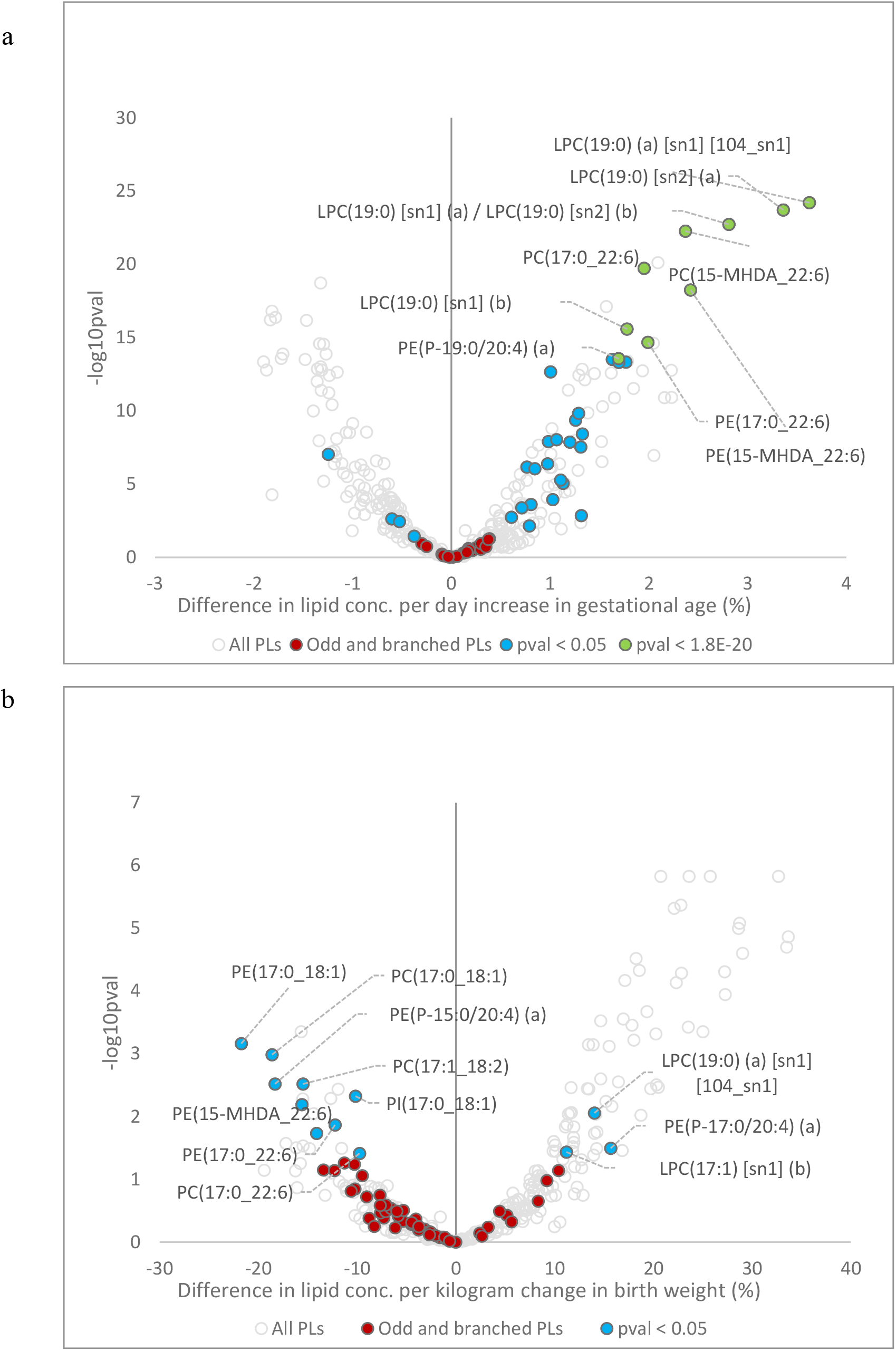
**a,** Volcano plot showing newborn phospholipids associated with gestational age. **b**, Volcano plot showing newborn plasma phospholipids associated with birth weight.

**Supplementary Fig 3.**
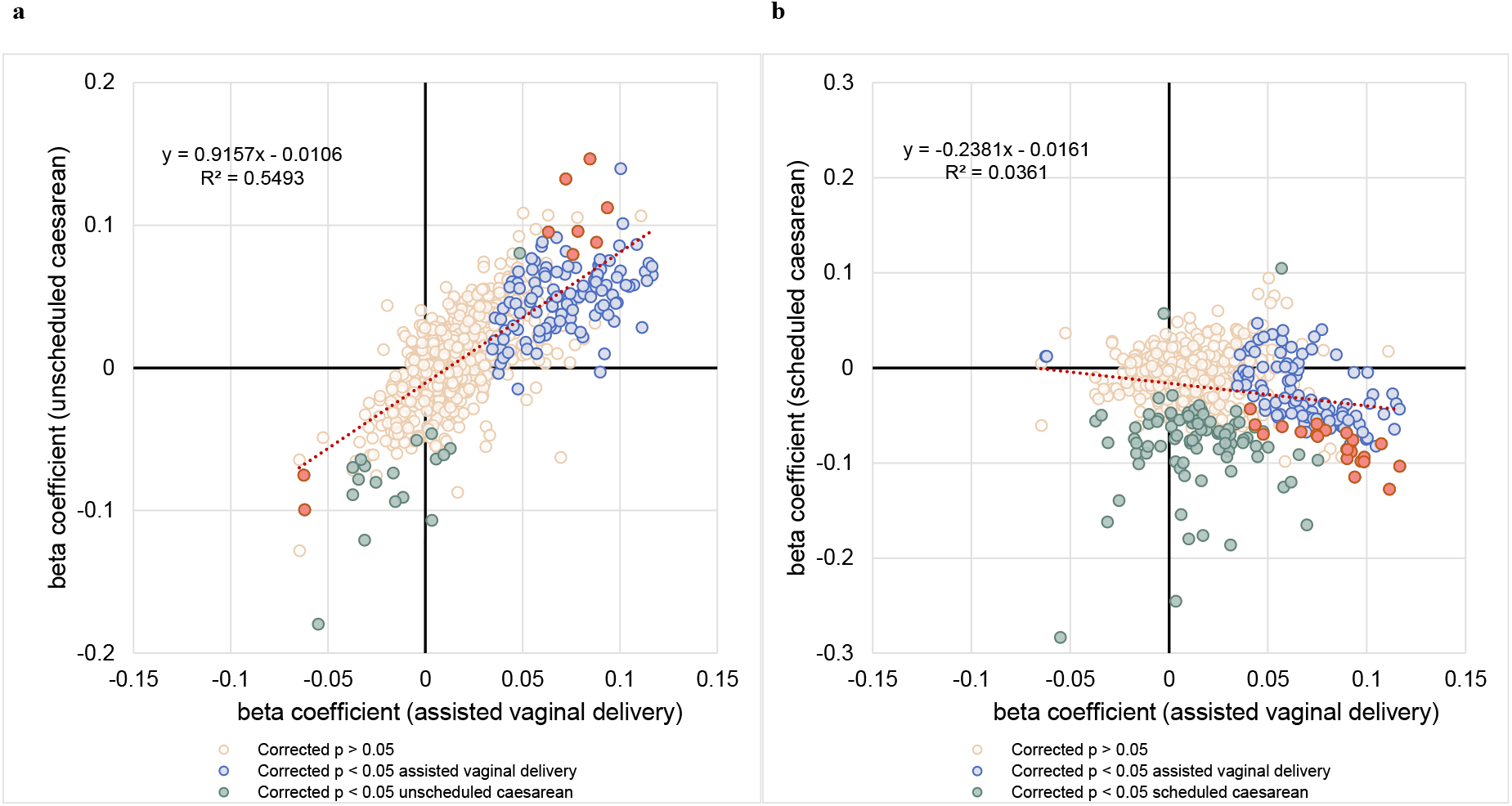
**a,** The correlation between regression coefficients of each lipid species associated with assisted vaginal delivery (x-axis) and lipid species associated with unscheduled cesarean delivery (y-axis) was examined. **b**, the correlation between regression coefficients of each lipid species associated with assisted vaginal delivery (x-axis) and lipid species associated with scheduled cesarean delivery (y-axis) was examined.

**Supplementary Fig 4.**
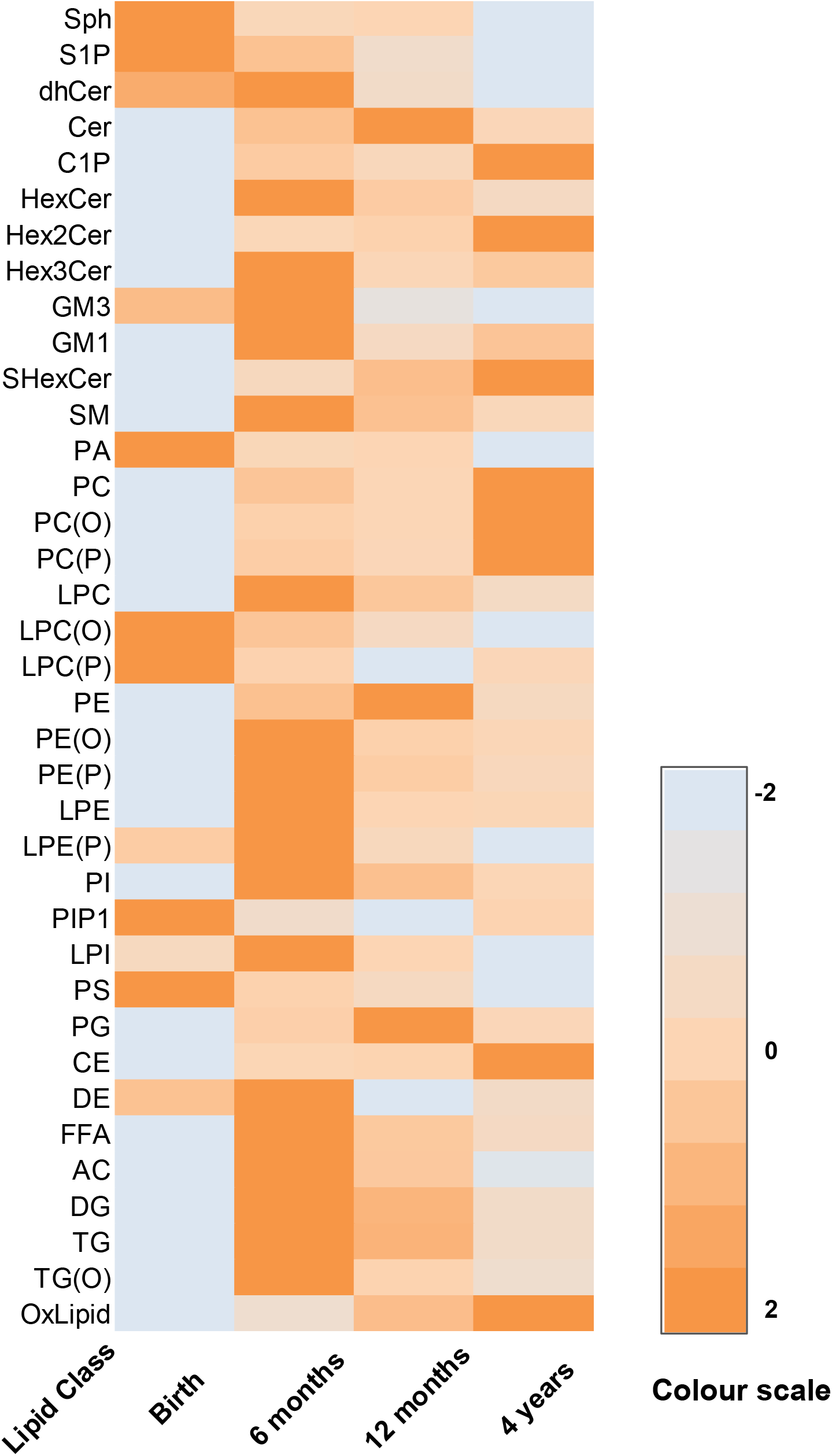
A heat map showing the concentration of lipid classes at each time point. Red: lowest value, blue: highest value

**Supplementary Fig.5.**
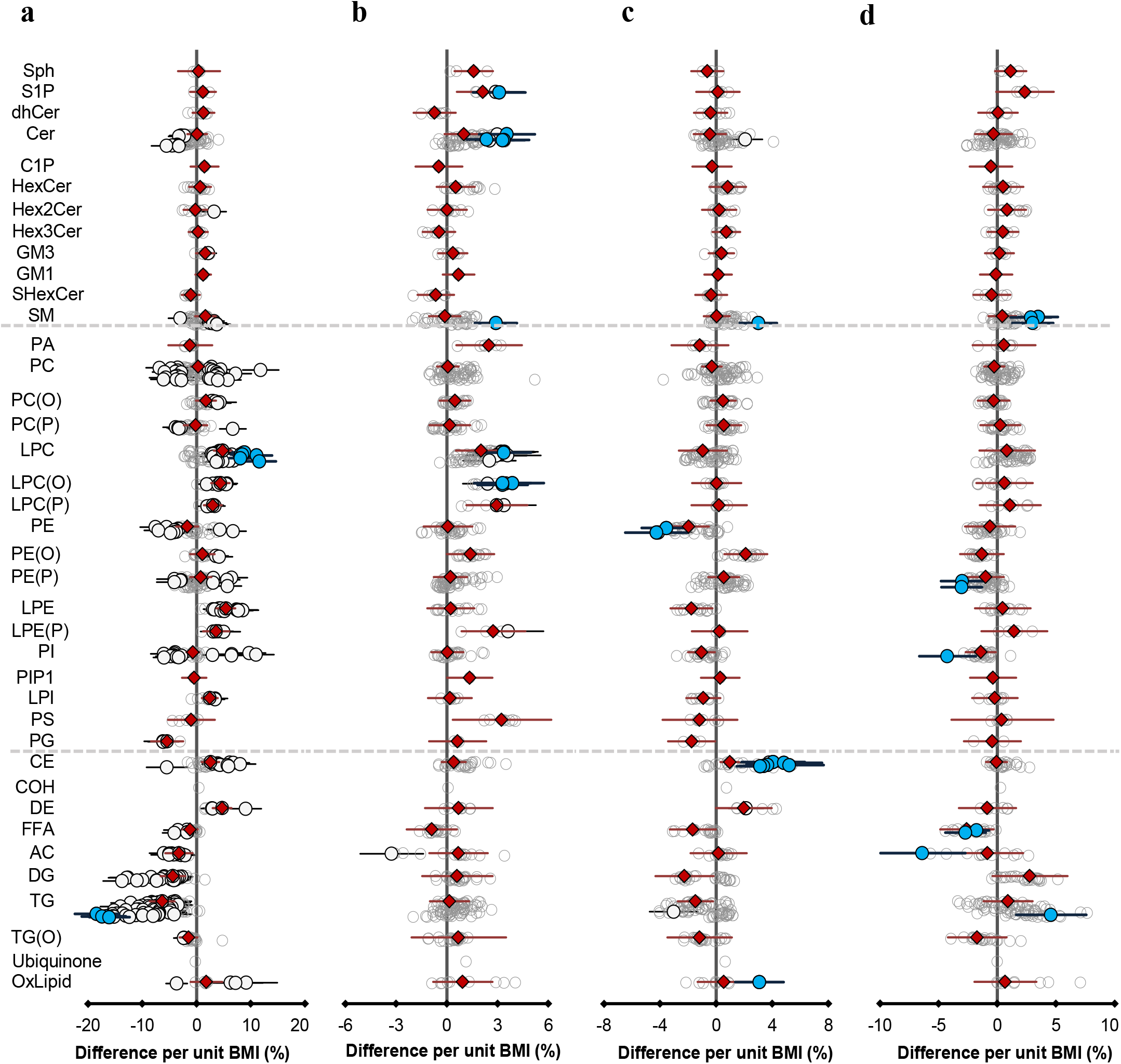
Association of infant BMI with lipid species. Estimated percentage difference in lipid concentrations per unit change in BMI at different time points, determined using linear regression analyses adjusting for age and sex and timepoint-specific covariates. **a**, Association of child BMI with cord serum lipidome at birth. **b**, Association of child BMI with plasma lipidome at six months. **c**, Association of child BMI with plasma lipidome at twelve months. **d**, Association of child BMI with plasma lipidome at four years. Results are shown as % difference in lipid concentration per unit change in BMI. Grey open circles show non-significant species, white closed circles show significant lipid species (p < 0.05), the top 10 lipid species are shown in blue p < 2.13E^-12^, 7.69E^-03^, 3.2E^-02^, 0.18red diamonds show lipid classes. All the p-values are after multiple testing comparison. Error bars represent 95% confidence intervals.

**Supplementary Fig 6.**
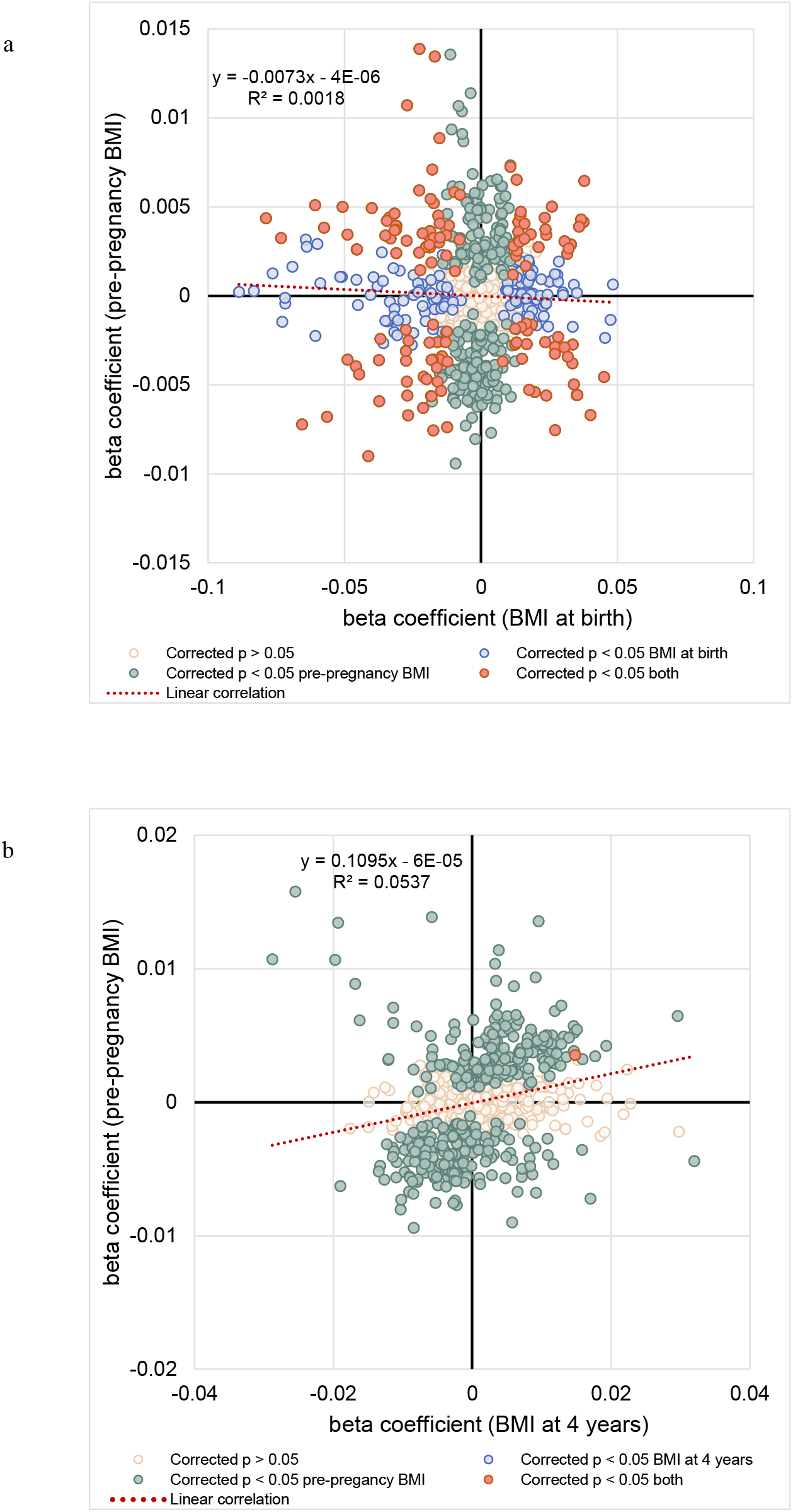
The correlation between regression coefficients of each lipid species associated with infant BMI and lipid species associated with adult BMI. **a,** The correlation between regression coefficients of each lipid species associated with BMI at birth (x-axis) and lipid species associated with pre-pregnancy BMI (y-axis) was examined. **b**, The correlation between regression coefficients of each lipid species associated with BMI at four years (x-axis) and lipid species associated with pre-pregnancy BMI (y-axis) was examined.

**Supplementary Fig 7.**
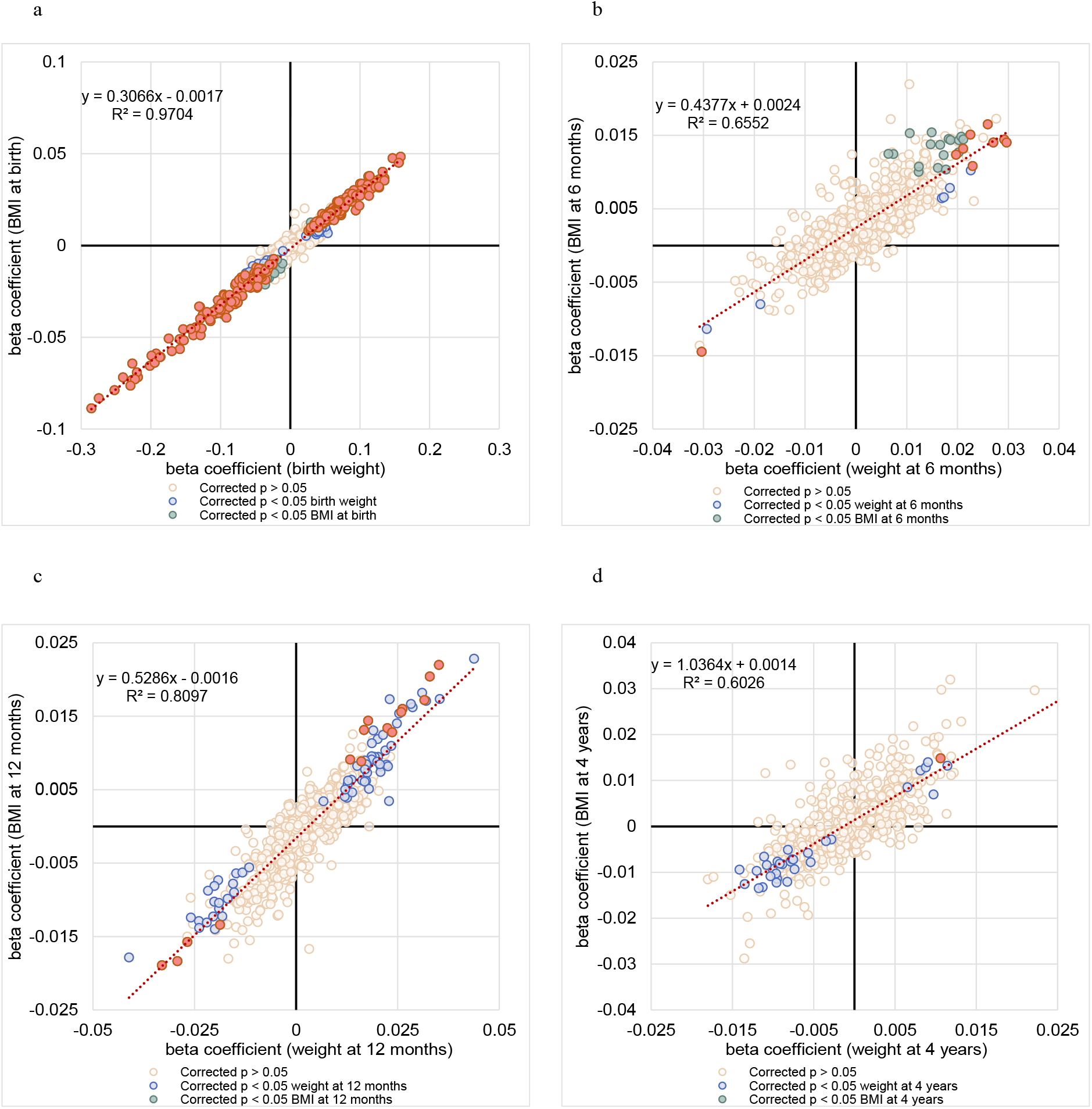
The correlation between regression coefficients of each lipid species associated with BMI and lipid species associated with weight at each time point. **a,** The correlation between regression coefficients of each lipid species associated with birth weight (x-axis) and lipid species associated with BMI at birth (y-axis) was examined. **b**, The correlation between regression coefficients of each lipid species associated with weight at 6 months (x-axis) and lipid species associated with BMI at 6 months (y-axis) was examined. **c**, The correlation between regression coefficients of each lipid species associated with weight at 12 months (x-axis) and lipid species associated with BMI at 12 months (y-axis) was examined. **d**, The correlation between regression coefficients of each lipid species associated with weight at four years (x-axis) and lipid species associated with BMI at four years (y-axis) was examined.

